# Artificial Intelligence-based Diagnosis of Kaposi Sarcoma using Photographs in Dark-skinned Patients

**DOI:** 10.1101/2025.04.21.25326060

**Authors:** Sarah J Coates, Feng Yang, Cody Hill, Zhiyun Xue, Sivaramakrishnan Rajaraman, Aggrey Semeere, Racheal Ayanga, Miriam Laker-Oketta, Helen Byakwaga, Robert Lukande, Matthew Semakadde, Micheal Kanyesigye, Megan Wenger, Philip LeBoit, Timothy McCalmont, Ethel Cesarman, David Erickson, Toby Maurer, Sameer Antani, Jeffrey Martin

## Abstract

**Importance:** Advanced-stage disease at the time of diagnosis, with resultant high mortality, is among the most urgent issues for HIV-related Kaposi sarcoma (KS) in sub-Saharan Africa. Lack of access to skilled clinical personnel and histopathologic technology in the region contribute to diagnostic delays and advanced stage at diagnosis. Accordingly, new paradigms for KS diagnosis are needed.

**Objective:** To evaluate the accuracy of artificial intelligence (AI)-based interpretation of digital surface images of skin lesions to diagnose KS among dark-skinned patients in Uganda.

**Design:** Cross-sectional study of consecutive participants referred to skin biopsy services in Uganda because of clinical suspicion of KS. Lesions were photographed using a digital camera, and punch biopsies were obtained. Histopathologic interpretation was considered the gold standard. Using training (∼70% of images) and validation (∼10% of images) sets, we developed a prediction model using a rule-based combination of YOLO (You Only Look Once) version 5 and 8 object detection classifiers.

**Setting:** Free-of-charge skin biopsy services

**Participants:** Consecutive sample of 482 individuals were evaluated due to clinical suspicion of KS.

**Main Outcomes:** Sensitivity, specificity, positive and negative predictive value (with accompanying 95% confidence intervals) of the AI-based prediction model in a test set (∼20% of images). The accuracy of a dermatologist’s visual interpretation of images was also described.

**Results:** 472 participants (1385 images) were evaluable. Of these, 36% were female, median age was 34 years, and 94% had HIV; 332 had KS, and 140 had no KS by histopathology. In the test set, the AI-derived prediction model achieved 89% sensitivity (85%-94%) and 51% specificity (40%-61%) for diagnosing KS; positive predictive value was 81% (75%-86%) and negative predictive value was 67% (55%-78%). A dermatologist evaluating the same images, with emphasis on sensitivity, achieved sensitivity of 93% (89%-96%) and specificity of 19% (11%-28%).

**Conclusions and Relevance:** Among dark-skinned patients in Uganda with skin lesions suspicious for KS, evaluation of digital surface images by an AI-based prediction model produced moderate accuracy for diagnosing KS. While currently inadequate for clinical use, this inaugural assessment is sufficiently promising to justify evaluation of larger datasets and evolving technologies to determine if accuracy can be improved.

**Key Points:** *Question:* Can an artificial intelligence (AI)-based prediction model be developed from digital images to accurately distinguish Kaposi sarcoma (KS) from non-KS in dark-skinned patients?

*Findings:* Evaluation of digital images of skin lesions from patients in Uganda by an AI-based prediction model produced moderate accuracy for diagnosing KS.

*Meaning:* In sub-Saharan Africa, where incidence and mortality from KS is high and delayed diagnosis is common due to limited specialized personnel and technical supplies, AI-based prediction models built on digital images taken of suspicious lesions may someday hasten KS diagnoses.

## Introduction

Even prior to the HIV epidemic, sub-Saharan Africa (SSA) had amongst the world’s highest incidence of Kaposi sarcoma (KS).^1^ Spread of HIV infection in the region subsequently transformed KS into a public health problem, becoming one of the most common^2,3^ and fatal cancers among adults.

While widespread antiretroviral therapy in SSA has begun to decrease KS incidence^4^ and improve survival, KS remains among the most commonly occurring cancers,^5^ and 1-year mortality among those with HIV-related KS has recently been estimated to be 41%.^6^ The most overt explanation for this poor survival is the high frequency of advanced-stage disease at the time of diagnosis.^7^ Many factors conspire for late-stage diagnosis of KS in SSA, including the inherent challenge of detecting early KS lesions in persons with dark skin types,^8^ lack of dermatologists^9^ and pathologists,^10^ and scarcity of technical materials (e.g., anti-latency-associated nuclear antigen staining) needed to assist in histopathologic diagnosis. Because the dearth of highly trained personnel and histopathologic technology shows no sign of abating soon, other innovative answers are urgently needed.

Automated interpretation of digital images of skin lesions suspicious for KS, a form of artificial intelligence (AI), is a possible solution to the paucity of human and technical resources needed to diagnose KS in SSA. Such an approach has the potential to place KS diagnostic capability into the hands of any clinician with a smartphone. Application of deep-learning (DL) algorithms has recently shown promise in the diagnosis of various skin cancers, including melanoma and keratinocyte carcinoma,^11^ as well as mucosal malignancies such as oral^12^ and cervical cancer.^13^ To our knowledge, this approach has not been evaluated for KS, nor has it been tested in any study population replete with dark-skinned individuals. Using photographs of skin lesions from patients in Uganda presenting with clinical suspicion of KS, we developed a DL-based prediction model and assessed its accuracy in diagnosing KS.

## Methods

### Overall Design

In a cross-sectional design, we obtained digital photographs of skin lesions from patients in Uganda referred to skin biopsy services because of clinical suspicion of KS. Biopsies of lesions were determined to be KS or not by U.S.-based pathologists, which was considered gold standard for the diagnosis. We assessed the accuracy for the diagnosis of KS of a DL-derived prediction model trained on visual features derived from the digital images paired with the histopathologic diagnosis. The accuracy of a dermatologist’s macroscopic interpretation of the images was also evaluated.

### Participants

We approached consecutive patients who were being evaluated because of clinical suspicion of KS by skin biopsy services at three medical centers in Kampala, Mbarara, and Masaka, Uganda. These skin biopsy services, which were staffed by a small group of providers who were originally trained by U.S.-based dermatologists, were established several years prior because of difficulty accessing histopathologic confirmation for KS in the region.^14^ The services provided punch biopsies performed on the same day as the initial visit free of charge to both outpatients and inpatients. Services were advertised to clinical providers throughout the respective local medical facilities, as well as facilities in the surrounding region, as a means to obtain a biopsy for patients with skin or mucosal lesions for whom the provider had suspicion of KS. Any outpatient who was able to travel to the biopsy venue was assessed. Inpatients from the respective medical centers were evaluated by the biopsy service team at the bedside. To our knowledge, these were the only free-of-charge skin biopsy services in Uganda at the time. Patients whose lesions were confirmed to be suspicious for KS by the biopsy team were offered a punch biopsy for histopathologic confirmation and asked if they wished to participate in our research study. Those who agreed provided written informed consent.

### Measurements

#### Questionnaire-based

Demographic, socioeconomic, and medical history information were collected on the day of biopsy.

#### Physical examination

A single lesion from each participant was selected to be biopsied and the biopsy provider recorded macroscopic morphology (macule/patch, plaque, papule/nodule or tumor).

#### Photographic

Each lesion was photographed with a digital camera (Canon EOS Rebel T6i DSLR or Canon Powershot ELPH 180). The team members had been briefly trained in a standardized approach to obtain images from multiple angles and varying distances from the lesion. For most lesions, three distinct images were taken. All images included rulers to record dimensions, and most included circular markings to denote the specific lesion biopsied.

#### Dermatologic

A U.S.-based board-certified dermatologist with specific expertise in HIV dermatology and experience practicing in sub-Saharan Africa interpreted all available digital images at the image level according to image quality (optimal, adequate, or inadequate) and, among those deemed optimal or adequate, they assessed the likelihood of KS: very unlikely KS (0-10% probability of KS); unlikely KS (11-30%); equivocal (31-70%); likely KS (71-90%); and very likely KS (91-100%). A second board-certified dermatologist with 25 years of experience also reviewed 20% of the images. Given a high level of agreement between the two dermatologists, the second dermatologist did not review the remaining images.

#### Histopathologic

Biopsies were processed, and a slide was stained with hematoxylin and eosin by a general pathology laboratory in Kampala, Uganda. In the U.S., an additional slide was stained with anti-latency-associated nuclear antigen, and both slides were used in the subsequent interpretation (KS, not KS, or indeterminate) by a KS-expert general pathologist and a dermatopathologist. If there was discordance between the two readers, a second dermatopathologist reviewed the case, and the majority interpretation was taken as the final diagnosis and the gold standard in subsequent analyses.

### Statistical Analysis

All images were included unless they were deemed inadequate by the dermatologist, defined as being unable to be evaluated by a human observer for the diagnosis of KS because the lesion biopsied was not apparent in the image or the image quality was too poor for assessment.

#### Development and evaluation of a digital image-derived DL-based prediction model for the diagnosis of KS

At the level of the participant, we first randomly split the photographs into training (∼70% of images), validation (∼10% of images), and hold-out test sets (∼20% of images). To account for potential differences in photographic technique and presence of KS across the three medical centers, the distribution of medical centers was kept equal across the three image sets. Each image was accompanied by its histopathology-derived gold standard diagnosis.

The prediction model was constructed, using the training and validation sets of images, as a two-tiered rule-based detection and classification system. A detector first learns to isolate and label all lesions in the entire image (Figure 1), and next, a classifier analyzes the highest-ranked single lesion identified by the first classifier. Because of imbalance in the number of KS and non-KS cases, several compensatory techniques were incorporated into the classifiers, including boosting minority class weights, focal loss that automatically down-weights the contribution of easy examples which are usually from the majority class and focuses on harder to classify samples,^15^ and data augmentation that increases the number of samples by inducing small variations in image appearance.^16^ We selected two versions of the You Only Look Once (YOLO) series^17^ of DL methods (YOLOv5^18^ and YOLOv8^19^), which are convolutional neural networks. We developed a YOLOv5-based lesion detection classifier to locate and label all skin lesions on the image, discarding all other image background, and labeling each lesion with a predicted probability of KS. We verified the lesion localization by randomly sampling YOLO output, resulting in displays of rectangular regions around the identified lesion (Figure 1). Within an image evaluated by YOLOv5, if no lesions were found, the image is labeled as “Non-KS” (Figure 2). If lesions are found, the demarcated lesion bounding box with the highest detection confidence is selected and its corresponding label is used to determine the image label (“KS” or “Non-KS”). The predicted lesion is automatically cropped from the image and classified using YOLOv8, and the resulting classification label is compared with the label from YOLOv5 in a rule-based decision. When both labels agree, the decision is sustained. In cases of disagreement, if YOLOv5 predicted the lesion as “Non-KS” while YOLOv8 predicted it as “KS”, then the image is labeled as “Non-KS”. This is because YOLOv5, in its role as an object detector, is considered superior to YOLOv8 lesion classification since, as part of its training, it has learned both – unique visual characteristics of the lesion and disambiguating them from other irrelevant image background. If YOLOv5 predicted it as “KS” with a confidence exceeding an empirically determined threshold of 0.6 but YOLOv8 disagreed, the image was still labeled as “KS”. This is because there are more KS samples in the training data and thus demanded the stricter threshold (greater than default of 0.5) which was determined using an internal validation set prior to final hold-out testing. Finally, the image-level prediction derived using this rule-based approach is compared to its respective accompanying gold standard histopathologic diagnosis. The performance was characterized using sensitivity, specificity, positive predictive value, and negative predictive value.

**Figure 1.**
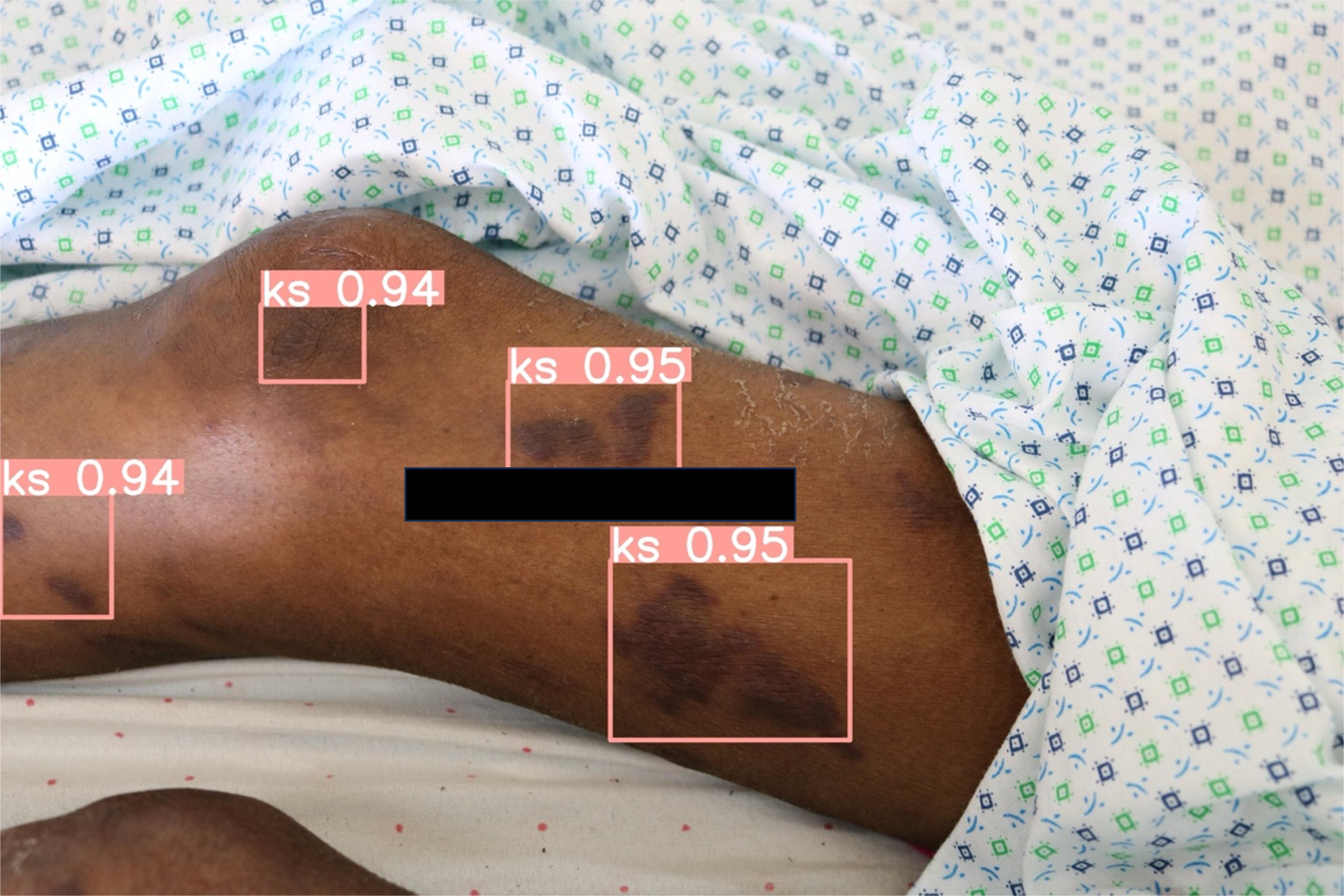
Digital image from representative participant illustrating lesion localization by YOLOv5 classifier, resulting in displays of rectangular regions around the identified lesion. Black rectangle is a redacted participant identification label.

**Figure 2.**
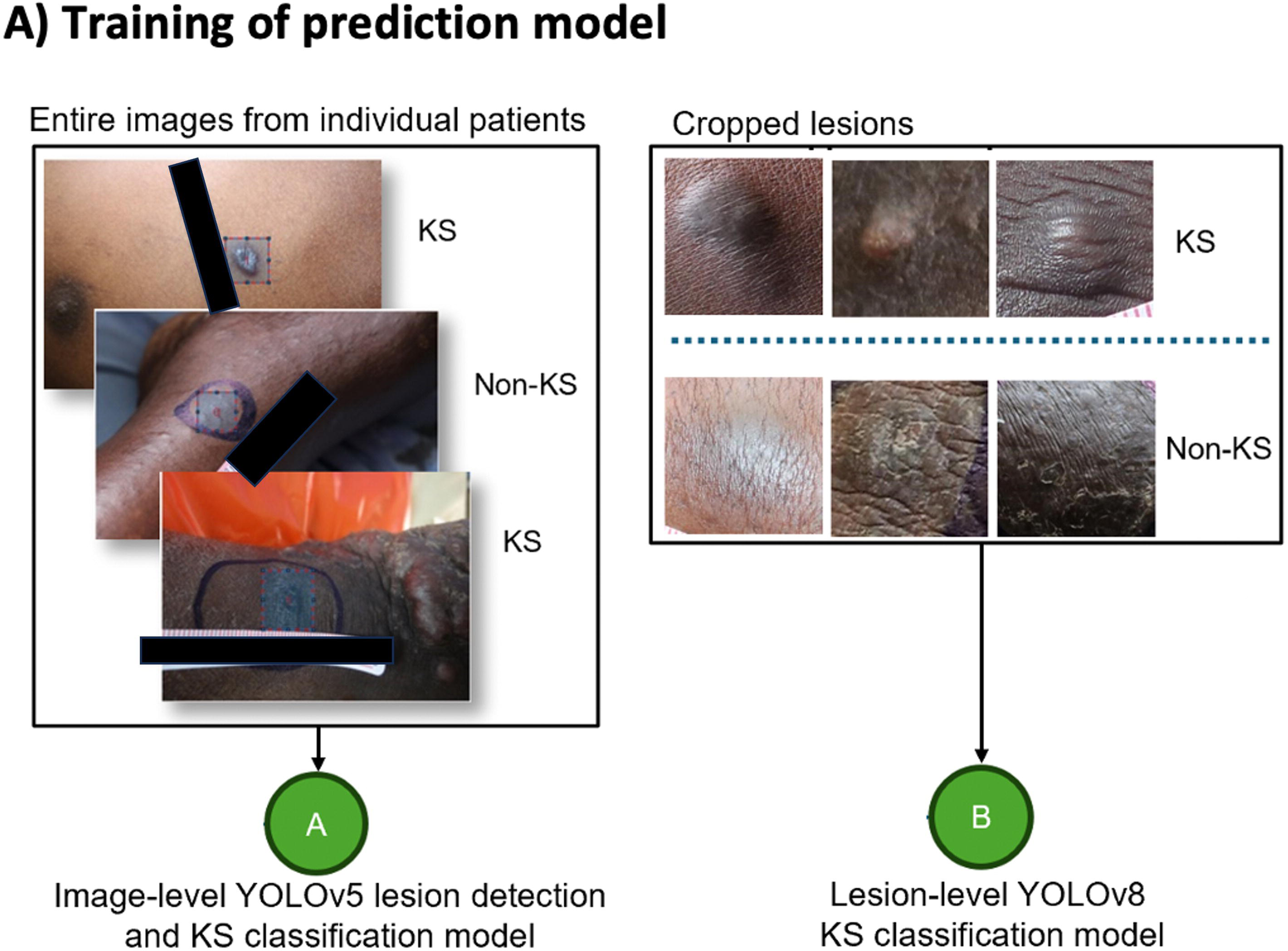
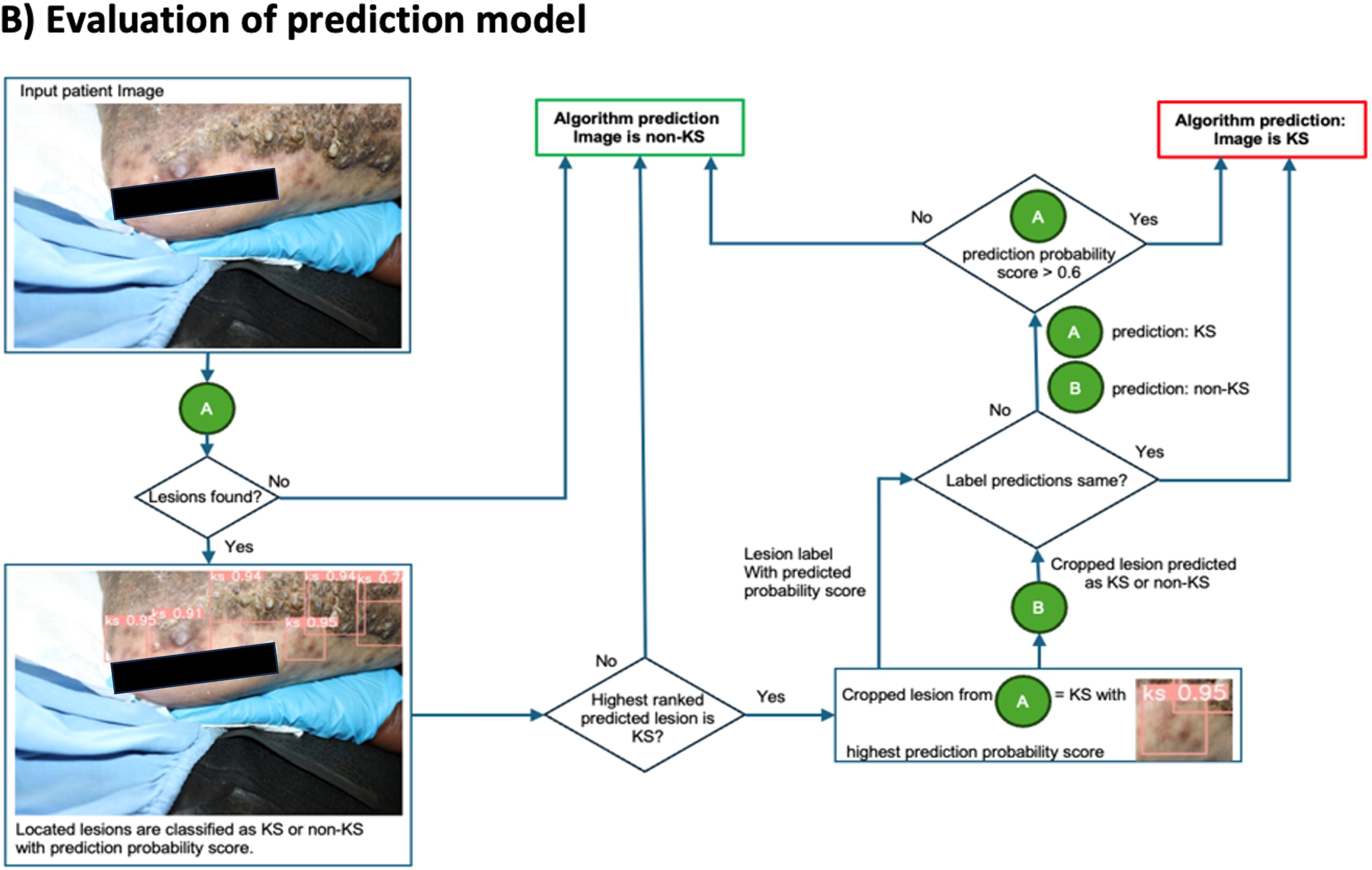
Description of the training and validation of the rule-based prediction model for the diagnosis of KS among participants who were referred to one of three free-of-charge skin biopsy services in Uganda because of clinical suspicion of KS. Panel A describes the two classifiers (YOLO version 5 and YOLO version 8) that are included in the prediction model. Panel B describes the rules used in determining the final classification of Kaposi sarcoma (KS) versus not KS by the prediction model.

#### Evaluation of a dermatologist’s macroscopic interpretation of digital images for the diagnosis of KS

To yield a binary classification, the dermatologist’s five-level interpretation of the probability of KS (very unlikely, unlikely, equivocal, likely, very likely KS) for each image was reduced to two levels (“KS” or “Non-KS”) using two different perspectives that depended upon the grouping of the equivocal level. In the first perspective, intended to enhance sensitivity, very likely, likely, and equivocal were considered as “KS” and the other two categories (very unlikely and unlikely) were “Non-KS”. In the second perspective, intended to enhance specificity, very likely and likely were considered as “KS”, and the other three categories as “Non-KS”. As described for the DL-based prediction model, the dermatologist’s interpretation was compared to the histopathologic gold standard.

## Results

### Characteristics of Participants

We initially evaluated 482 participants. After removing images with inadequate quality, the dataset consisted of 472 participants (1385 images); 36% were female, the median age was 34 years, and 94% were living with HIV (Table 1). There was a wide distribution of anatomic skin sites biopsied as well as macroscopic morphology of the biopsied lesion. As evidence of the lack of ready access to biopsy services in the region, the median one-way travel time for the participant to the biopsy facility was one hour at a median cost of nearly $3 U.S. dollars. For context, this is 4% of the mean monthly income in Uganda ($81 U.S. dollars).^20^ Histopathologic interpretation of the lesions biopsied in these participants revealed that 332 had KS and 140 did not have KS.

**Table 1.**
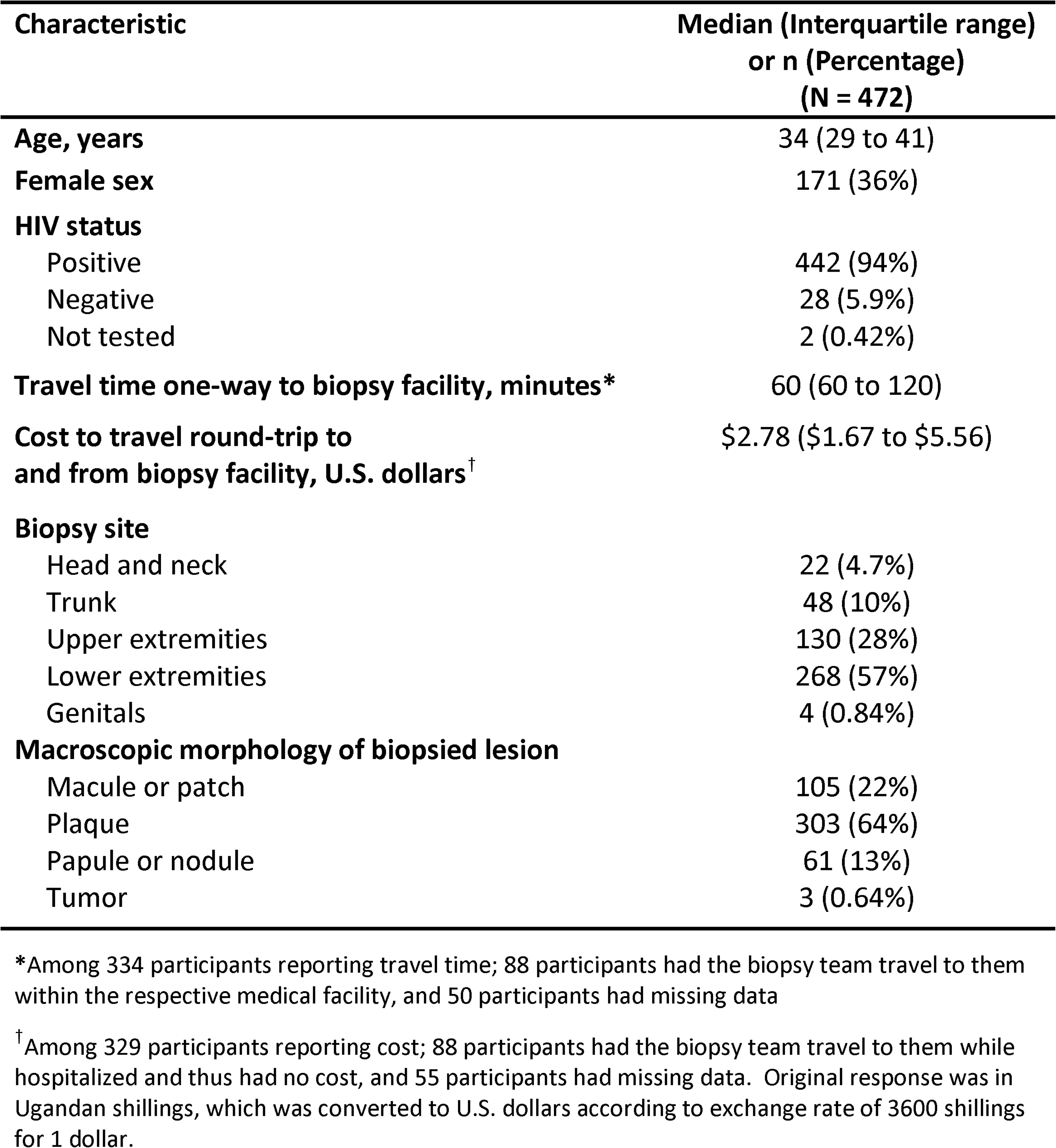
Characteristics of participants who were referred to one of three free-of-charge skin biopsy services in Uganda because of clinical suspicion of KS and who had a biopsy procedure performed.

| Characteristic | Median (Interquartile range)<br>or n (Percentage)<br>(N = 472) |
| --- | --- |
| Age, years | 34 (29 to 41) |
| Female sex | 171 (36%) |
| HIV status |  |
| Positive | 442 (94%) |
| Negative | 28 (5.9%) |
| Not tested | 2 (0.42%) |
| Travel time one-way to biopsy facility, minutes* | 60 (60 to 120) |
| Cost to travel round-trip to<br>and from biopsy facility, U.S. dollars<sup>†</sup> | \$2.78 (\$1.67 to \$5.56) |
| Biopsy site |  |
| Head and neck | 22 (4.7%) |
| Trunk | 48 (10%) |
| Upper extremities | 130 (28%) |
| Lower extremities | 268 (57%) |
| Genitals | 4 (0.84%) |
| Macroscopic morphology of biopsied lesion |  |
| Macule or patch | 105 (22%) |
| Plaque | 303 (64%) |
| Papule or nodule | 61 (13%) |
| Tumor | 3 (0.64%) |

\*Among 334 participants reporting travel time; 88 participants had the biopsy team travel to them within the respective medical facility, and 50 participants had missing data
<sup>†</sup>Among 329 participants reporting cost; 88 participants had the biopsy team travel to them while hospitalized and thus had no cost, and 55 participants had missing data. Original response was in Ugandan shillings, which was converted to U.S. dollars according to exchange rate of 3600 shillings for 1 dollar.

### Accuracy of the DL-derived Prediction Model in Diagnosing KS

The training set was allocated approximately 70% (970 images) of the images, the validation set 10% (140 images) of the images, and the test set 20% (275 images). After development and fine-tuning of the prediction model using the training and validation set of images, we evaluated the model in the test set of 275 images, which was comprised of 66 participants with KS and 29 without KS (Figure 3). In the test set, the prediction model had a sensitivity of 89% (95% confidence interval (CI): 85% to 94%) and specificity of 51% (95% CI: 40% to 61%) for the diagnosis of KS. Positive predictive value was 81% (95% CI: 75% to 86%), and negative predictive value was 67% (95% CI: 55% to 78%) (Table 2).

**Figure 3.**
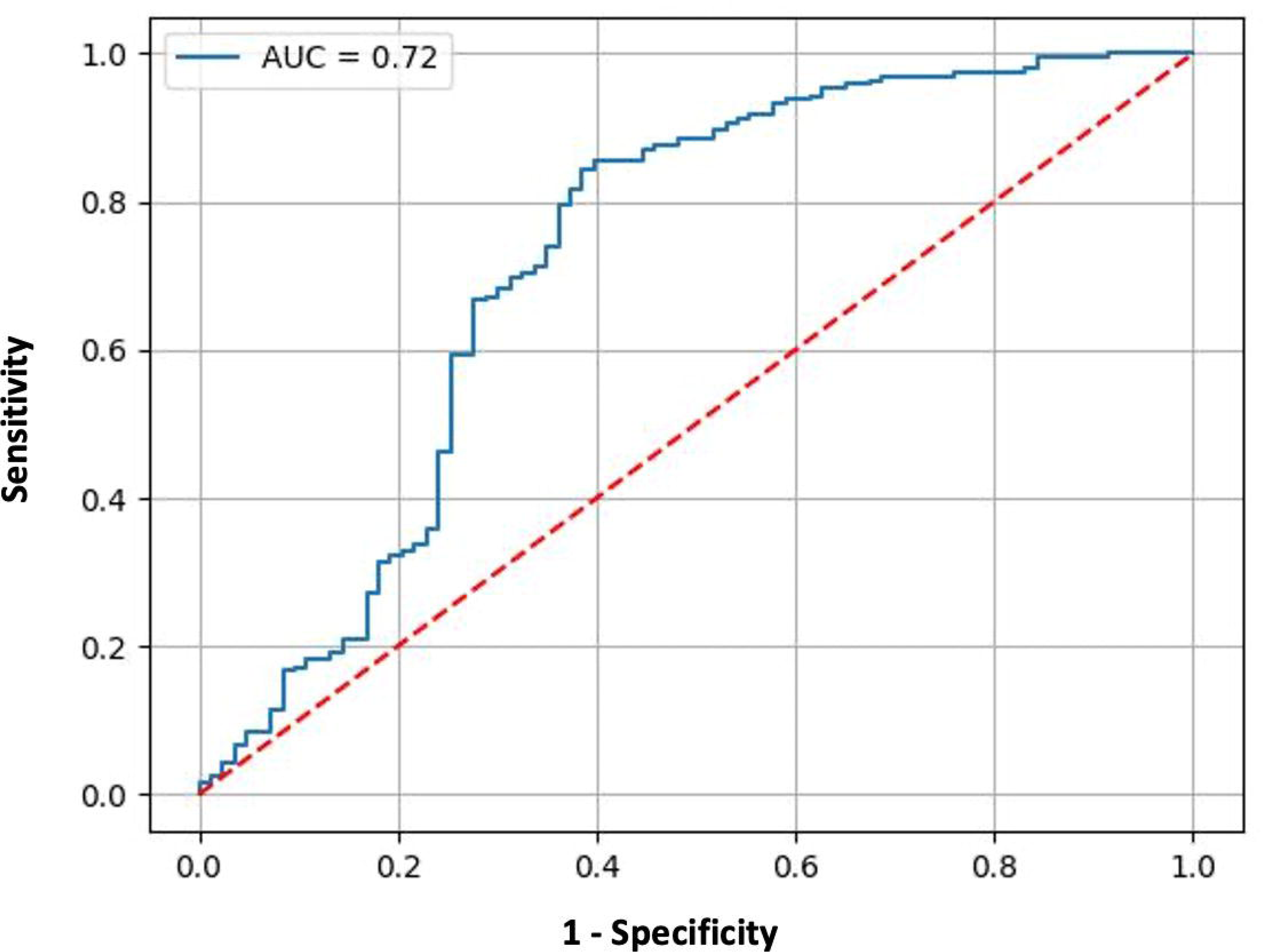
Receiver operating characteristics (ROC) curve depicting the performance of the digital image-based prediction model for the diagnosis of Kaposi sarcoma (KS) among 275 images in the test set derived from consecutive participants who were referred to one of three biopsy services in Uganda because of clinical suspicion of KS.

**Table 2.**
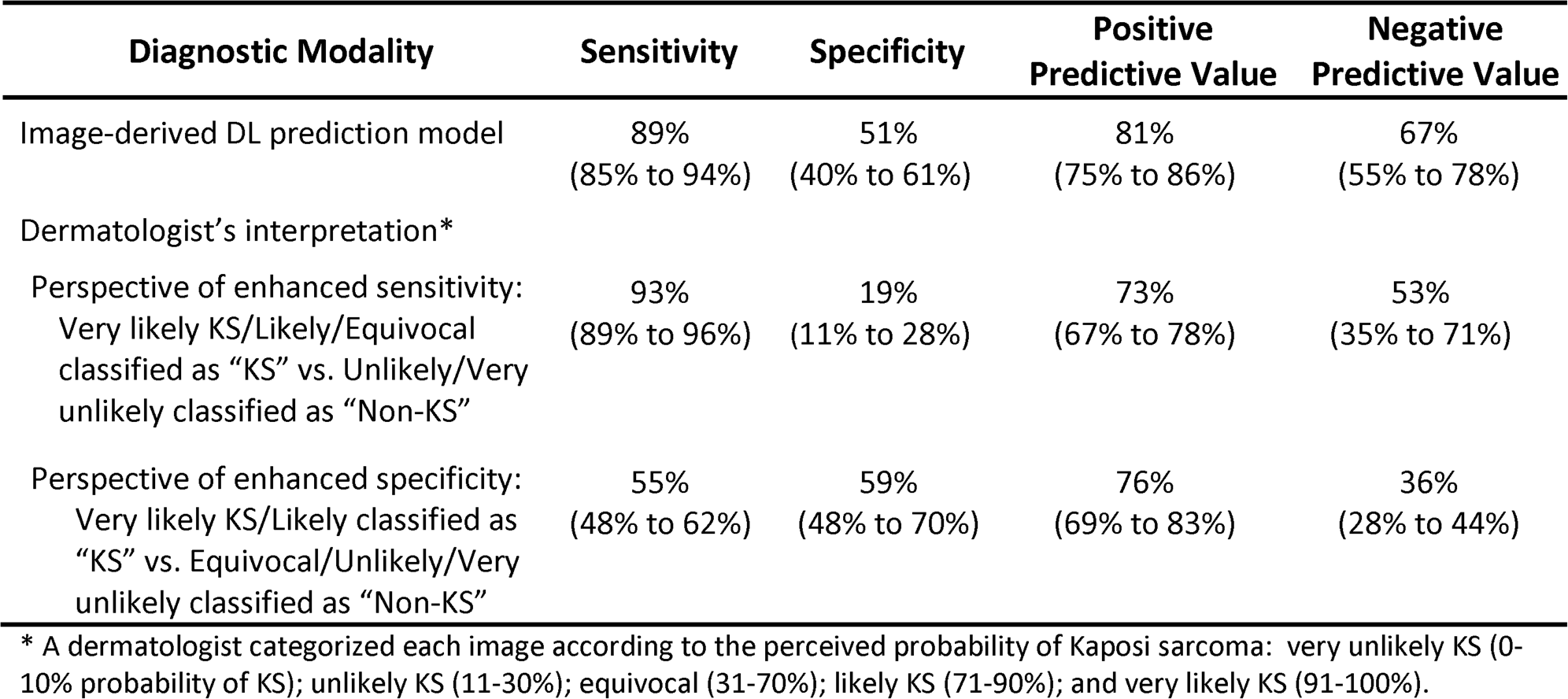
Image-level performance estimates and 95% confidence intervals on the test set of 275 images of a) a rule-based combination of YOLOv5 and YOLOv8 predictions using digital images of skin lesions; and b) a dermatologist’s macroscopic interpretation of the digital images (two perspectives were examined). The test set is a 20% sample of images taken of skin lesions from participants who were referred to one of three skin biopsy services in Uganda because of clinical suspicion of Kaposi sarcoma.

### Performance of the Dermatologist in Diagnosing KS

During the initial phase of the study, when the dermatologist interpreted the macroscopic appearance of all 1385 images, 12% were classified as very likely KS, 44% as likely, 34% as equivocal, 9.6% as unlikely, and 0.43% as very unlikely. To yield a binary classification from the dermatologist’s interpretation (to contrast with binary classification from the DL-based prediction model), images labeled as equivocal needed to be grouped either with the “KS” or “Non-KS” category. In a perspective aimed towards enhancing sensitivity, equivocal images were classified as “KS”. In a perspective to enhance specificity, equivocal images were classified as “Non-KS”. Among the test set of 275 images, in the perspective that enhanced sensitivity, the dermatologist’s interpretation yielded a sensitivity of 93%, specificity of 19%, positive predictive value of 73%, and negative predictive value of 53% (Table 2). In the perspective that enhanced specificity, sensitivity was 55%, specificity 59%, positive predictive value 76%, and negative predictive value 36%. In the entire set of 1385 images, the dermatologist’s accuracy was qualitatively similar to the test set. In the perspective that enhanced sensitivity, sensitivity was 95%, specificity 22%, positive predictive value 75%, and negative predictive value 63%. In the perspective that enhanced specificity, sensitivity was 65%, specificity 67%, positive predictive value 83%, and negative predictive value 44%.

## Discussion

In SSA, despite widespread availability of antiretroviral therapy in the “Treat All” era, KS remains among the most common^2,3^ and fatal cancers.^6^ Many factors combine to result in poor survival after KS diagnosis in the region, but the high frequency of advanced-stage disease at diagnosis is among the most prominent.^7^ Why patients are not formally diagnosed earlier has many explanations, but difficulty accessing histopathologic diagnosis is clearly a root cause. As such, innovation in KS diagnosis is needed. To address this, we developed and assessed the diagnostic accuracy of a DL-based prediction model trained on the visual features of digital images of skin lesions obtained from dark-skinned patients with clinical suspicion for KS. While we found that the prediction model had moderate accuracy for the diagnosis of KS, and is currently inadequate for clinical use, this inaugural assessment is sufficiently promising to justify further work.

There is no prior research evaluating the interrogation of skin surface digital images for the diagnosis of KS, but it is interesting to compare our findings to the use of digital images to diagnose other skin diseases. Our model for KS has a lower sensitivity (89%) and specificity (51%) than models diagnosing melanoma and keratinocyte carcinoma, for which the most recent studies have yielded sensitivity above 90% and specificity above 80%.^21,22^ There are several potential explanations for this difference. First, we had fewer images in our training dataset compared to other studies,^22^ and it is conceivable using more images to train could produce greater accuracy. Second, KS may be inherently more difficult to distinguish from its mimickers. KS lesions have several morphologies (e.g. patch, plaque, nodule) as well as a wide between-person distribution of number of lesions and anatomic regions involved. As such, there are many conditions that mimic KS, making it more difficult to distinguish KS from its mimickers. This is distinct, for example, from melanoma, in which the question for prediction model is primarily melanoma versus one mimicker, benign melanocytic nevus. Third, it may be that distinguishing skin conditions by analysis of surface morphology is intrinsically more challenging in dark-skinned patients than fair-skinned persons in whom most of the initial AI-related work in dermatologic diagnosis has thus far been performed. Only more research in dark-skinned patients will answer this possibility.

We believe our work is also the first to evaluate the accuracy of a dermatologist in diagnosing KS, and several findings are notable. First, about one third of images were interpreted by the dermatologist as equivocal for the diagnosis of KS. Second, regardless of how the equivocal images were classified in our analyses, the resultant accuracy is insufficient to either rule in or rule out KS. Collectively, these findings provide evidence for what is generally believed by dermatologists, that KS is difficult to diagnose on clinical grounds alone and thus an objective means of diagnosis is needed (which heretofore has been histopathology) when there is any level of clinical suspicion.

We might have benefited from augmenting our dataset with images from online databases, and indeed attempted to do so, but there is a paucity of images from dark-skinned patients in these resources. Widely used histopathology-confirmed databases, such as the International Skin Imaging Collaboration, include images mostly from countries with fair-skinned populations.^23^ Among machine-learning studies focused on diagnosis of skin cancers, the majority do not disclose skin types of image sources, and of those that do, very few include patients with dark skin types.^24–26^ In available databases, there is especially a paucity of dark-skinned images that have associated histopathological confirmation; this reflects the practice settings of most dermatologists worldwide, disparate access to dermatology care in racially diverse countries, and the systems that are in place to deliver healthcare — including pathology services — to dark-skinned patients throughout the world.^27^ This has implications for the accuracy of AI developed from currently available resources. In Uganda, the AI dermatological algorithm Skin Image Search was substantially less accurate when provided images of conditions in people with darker skin colors.^28^ Daneshjou et al. created the Diverse Dermatology Images (DDI) data set, “the first publicly available, expertly curated, pathologically confirmed image data set with diverse skin tones”, and showed that current state-of-the-art dermatology AI models were less accurate in patients with dark skin tones.^27^ This emphasizes the need to continue advancing representation of dark skin types in this space.

There are several limitations of our work. First, the context of our biopsy service meant only lesions that were suspicious for KS from the standpoint of the biopsy service team were biopsied. There were likely some instances of non-KS that were excluded that would not have been excluded by less experienced practitioners in real-world settings. Because we do not know the prevalence of such lesions in current clinical practice, it is unclear how their presence would alter the diagnostic accuracy of our prediction model. Second, it is conceivable that diagnostic accuracy of image interpretation might have been better if we had required standardized image acquisition using professional photographers. We chose not to pursue this and instead focused on more practical photography that was realistic in real-world settings. Third, as mentioned earlier, we had fewer images to train on compared to earlier studies of other skin cancers. A larger number of images might have improved accuracy. Finally, given that this model was trained exclusively using dark-skinned persons, our results are not necessarily generalizable to all patients with lesions concerning for KS.

Although our AI prediction model generally performed better than a dermatologist, the international standard for KS diagnosis is histopathologic interpretation (for which accuracy is implied to be 100% sensitive and specific). Accordingly, in comparison to histopathology, our prediction model in its current form is not sufficiently accurate for clinical use. Nonetheless, even if continued enhancement of the model fails to make it equivalent to histopathology, there may be utility of even an imperfect prediction model. First, a model with very high sensitivity and acceptable specificity could be useful in triaging lesions initially being evaluated by a non-dermatologist provider and reduce some unnecessary biopsies. Second, a model with high specificity could be used to diagnose the presence of KS in some fraction of patients, thereby hastening subsequent management. Of note, if and when a clinical role is identified for this prediction model in dark-skinned patients in SSA, there may also be a role in the U.S., where black men with HIV infection represent the only group who are currently experiencing increases in KS incidence.^29^

In summary, among dark-skinned patients in Uganda with skin lesions clinically suspicious for KS, a DL-based prediction model trained on digital images of the lesions had moderate accuracy in diagnosing KS. The diagnostic accuracy of the prediction model appeared to be superior to a dermatologist, but the diagnostic performance of the model in its current state is insufficient for clinical use. Nonetheless, this inaugural assessment is sufficiently promising to justify further evaluation of larger image sets and evolving DL technologies to determine if accuracy can be improved.

## Data Availability

All data produced in the present study are available upon reasonable request to the authors

## Acknowledgments

This research was supported by the National Institutes of Health (UH2 CA202723, UH3 CA202723, U54 CA190153, U54 CA254571, U01 CA269199, D43 TW009343, P30 AI027763, and the Intramural Research Program of the National Library of Medicine) and the University of California Global Health Institute (UCGHI). The content is solely the responsibility of the authors and does not necessarily represent the official views of the NIH or UCGHI.

